# Utilization of Epigenome-wide DNA Methylation for Longitudinal Comparison of White Blood Cell Proportions Across Preeclamptic and Normotensive Pregnancy by Self-Reported Race

**DOI:** 10.1101/2020.09.18.20197491

**Authors:** Mitali Ray, Lacey W. Heinsberg, Yvette P. Conley, James M. Roberts, Arun Jeyabalan, Carl A. Hubel, Daniel E. Weeks, Mandy J. Schmella

## Abstract

**Objective:** We utilized epigenome-wide DNA methylation data to estimate/compare white blood cell (WBC) proportions in plasma across preeclamptic (case) and uncomplicated, normotensive (control) pregnancy.

**Methods:** We previously collected methylation data using Infinium® MethylationEPIC Beadchips during the three trimesters in 28 cases and 28 controls (21 Black, 7 White participants/group). We employed Houseman’s regression calibration method to estimate and compare neutrophil, monocyte, B cell, NK cell, CD4+ T and CD8+ T cell proportions across pregnancy and between cases and controls.

**Results:** We observed changes in WBC proportions across pregnancy within cases and controls that varied by cell type and race. Neutrophils represented the largest WBC mean proportion in all three trimesters for cases (Mean±SD: 67.2±9.6% – 74.4±12%) and controls (64.2±11% – 74.0±7.9%). Mean B cell proportions were significantly lower in cases than controls in Trimester 1 (5.25±0.02% versus 6.30±0.02%, *p*=0.02). The remaining mean cell proportions did not significantly differ in the overall sample. Stratified analyses revealed race-specific differences. In White participants (n=14): (1) neutrophil proportions were significantly higher in cases in Trimester 1 (*p*=0.04), but significantly lower in Trimester 2 (*p*=0.02), (2) B cell proportions were significantly lower in cases in Trimester 1 (*p*=0.001). No significant differences were detected among Black participants (n=42).

**Conclusions:** Although chronic inflammation characterizes preeclampsia, few studies have investigated WBCs across pregnancy. We report differences between cases and controls across pregnancy. Our findings in a small sample demonstrate the need for additional studies investigating the relationship between race and WBCs in pregnancy, which could provide insight into preeclampsia pathophysiology.

## Introduction

Preeclampsia (PE) is a leading cause of maternal and prenatal morbidity, affecting 2.3 to 3.8% of pregnancies in the United States, and 4.6% of pregnancies globally (Abalos, Cuesta, Grosso, Chou, & Say, 2013; Ananth, Keyes, & Wapner, 2013). Furthermore, individuals who develop PE are 3-4 times more likely to develop hypertension and have double the risk for experiencing a myocardial infarction or stroke later in life (Valdes, 2017). There are presently no therapeutic agents available for individuals who develop PE; the sole treatment is delivery of the fetus and placenta, which can result in morbidity or mortality of the fetus, depending upon the gestational week at which the fetus is delivered (Chappell et al., 2019; Coviello et al., 2019; Pauli & Repke, 2015).

The lack of available treatment options for PE patients is a direct consequence of our ongoing, limited understanding of its pathophysiology. PE development highlights the intersection between inflammation and angiogenesis, two directly linked biological processes that share several factors (Agarwal & Karumanchi, 2011; Catarino et al., 2012; Costa, Incio, & Soares, 2007; Escudero, Roberts, Myatt, & Feoktistov, 2014; Harmon et al., 2016; Jackson, Seed, Kircher, Willoughby, & Winkler, 1997). One potential mechanism of PE proposed in the literature is that inappropriate angiogenesis results in hypoxia, which drives inflammation, fueling a vicious cycle of chronic inflammation, dysregulated angiogenesis, hypoxia or ischemia-reperfusion injury, and systemic endothelial dysfunction (Amaral, Wallace, Owens, & LaMarca, 2017; Rana, Lemoine, Granger, & Karumanchi, 2019; Shamshirsaz, Paidas, & Krikun, 2012).

Despite decades of research, the exact mechanisms of PE pathophysiology remain elusive and while potential biomarkers have been identified, none have been translated into present routine clinical care (Anderson & Schmella, 2017; Polsani, Phipps, & Jim, 2013; Sunjaya & Sunjaya, 2019).

A major obstacle in PE research is limited access to samples. The majority of studies investigating PE focuses primarily on the placenta (Nelson, Ziadie, McIntire, Rogers, & Leveno, 2014; Staff et al., 2013; Vinnars et al., 2008). While these data are important for elucidating PE pathophysiology, one drawback of solely studying the delivered placenta is that it severely limits the scope of PE research by only providing one snapshot in time, specifically when the disease has become clinically overt and requires intervention. It is not possible to access the placenta throughout pregnancy ethically, unless the individual requires sampling for another medically indicated reason. Additionally, there is a need for establishment of an early biomarker, which requires investigation of easily accessible, non-invasive tissue. Lack of access to the placenta obstructs early identification of those at highest risk, which further prevents surveillance.

Peripheral blood represents a minimally invasive sample that can be collected at multiple points in time, allowing for longitudinal analysis, and therefore providing multiple snapshots. A reliable early biomarker would allow for earlier diagnosis of PE and closer surveillance throughout pregnancy.

White blood cells (WBCs) are known to be elevated in individuals across pregnancy (Abbassi-Ghanavati, Greer, & Cunningham, 2009; Gabbe, 2017; Pitkin & Witte, 1979). It is important to note that WBC values vary across race in healthy people and as such, it is possible for changes in WBCs during pregnancy to also differ with race (Lim, Cembrowski, Cembrowski, & Clarke, 2010). Moreover, few studies have investigated the differences in specific WBC values and phenotypes in individuals who develop PE compared to normotensive individuals, a significant gap in PE research (Kirbas et al., 2015; Lurie, Frenkel, & Tuvbin, 1998; Örgül et al., 2019). Furthermore, a limitation of these studies is that they are cross-sectional. Longitudinal quantification of potential changes in WBCs across pregnancy in individuals who develop PE represents a gap in the literature we sought to fill.

Typically, quantification of WBCs for biological research is performed by flow cytometry, which requires immediate sample preparation and assessment. Researchers who have approved access to clinical laboratory values, including a complete blood count with differential, may collect these data from patient charts. Another method that may be utilized by researchers is the Houseman approach, which allows for WBC proportions to be inferred from differentially methylated regions that serve as unique signatures for specific cell types (Houseman et al., 2012). In cases where epigenome-wide DNA methylation data exist, researchers have the opportunity to capitalize on these data to evaluate WBC proportions without using any additional resources. To date, we have not identified any studies that have published such estimated WBC proportions for PE cases versus normotensive controls. The purpose of this study was to characterize WBC proportions estimated from epigenome-wide DNA methylation data collected across the three trimesters of pregnancy in individuals who developed PE (cases) and individuals who remained normotensive throughout their uncomplicated pregnancy (controls). We hypothesized that WBC proportions for PE cases would differ from normotensive controls, reflecting a chronic inflammatory state.

## Methods

### Participants and Phenotype Designations

Participants included in this study were enrolled in a prospective cohort-based study funded by the National Institute of Child Health and Human Development (NICHD), entitled “Prenatal Exposures & Preeclampsia Prevention Project 3 (PEPP3): Mechanisms of Preeclampsia and the Impact of Obesity” [P01 HD30367]. The purpose of PEPP3, which took place at University of Pittsburgh Medical Center Magee-Womens Hospital from 2008 – 2014, was to explore factors associated with PE and obesity. As such, there was an emphasis on the recruitment of overweight and obese participants (BMI ≥ 25 kg/m^2^).

The current study consisted of n=28 participants diagnosed with PE (cases) and n=28 control participants who were 1:1 frequency matched on pre-pregnancy BMI, gestational age at sample collection, self-reported race, and self-reported smoking status (defined by <100 lifetime cigarettes). Pregnancy outcomes were based on a thorough review of clinical data by an expert panel of clinicians and researchers. Individuals who were <14 or >40 years of age, had a multiple pregnancy, or had a past medical history of conditions associated with heightened risk for PE (e.g., chronic hypertension, chronic renal disease, diabetes) were excluded from the study. We defined PE as new onset of gestational hypertension in previously normotensive individuals, after gestational week 20, and the presence of proteinuria (ACOG, 2002). Gestational hypertension was defined as new onset of elevated blood pressure (systolic BP ≥ 140 mmHg and/or diastolic BP ≥ 90 mmHg) that reverted to baseline by 12 weeks postpartum. The mean of the previous four blood pressures measured in labor and delivery, prior to any therapeutic intervention, was compared to mean blood pressures determined before 20 weeks’ gestation.

Proteinuria was defined as ≥ 300 mg over 24 hours, ≥ 0.3 protein/creatinine ratio, ≥ 2+ on a random urine dipstick, or ≥ 1+ on a catheterized urine specimen. Participants who served as the control phenotype were individuals who remained normotensive throughout the entirety of pregnancy, did not develop proteinuria, and delivered a healthy infant.

### Biospecimen Sampling, DNA Extraction, and DNA Methylation Data Collection

A peripheral blood sample was collected in EDTA plasma tubes from each participant in the first, second, and third trimester of pregnancy (one sample/trimester). For each sample, WBCs were isolated, and nuclear DNA was extracted, using protein precipitation methods.

Epigenome-wide DNA methylation data were collected using the Infinium MethylationEPIC Beadchip (Illumina, San Diego, CA, USA) at the Johns Hopkins University Genetic Resources Core Facility SNP Center (data available from dbGAP, accession number: phs001937.v1.p1). Twenty-eight technical replicates were included, and to mitigate potential batch effects, we attempted to place all samples from each participant on the same chip. To balance potential row and column effects, our sample layouts had an equal number of cases and controls in each of the two columns of each chip and a zigzag pattern was utilized from row to row, alternating whether a case sample or a control sample was in the first row of the chip.

Our epigenome-wide DNA methylation data cleaning and quality control pipeline was carried out in R using the minfi, ENmix, and funtooNorm packages (Aryee et al., 2014; Oros Klein et al., 2016; Xu, Niu, Li, & Taylor, 2016). This pipeline included removal of poorly performing and outlying samples and probes, as well as functional normalization (Aryee et al., 2014; Oros Klein et al., 2016; Xu, Langie, De Boever, Taylor, & Niu, 2017; Xu et al., 2016). Our final data set consisted of 703,200 probes available in 56 participants at up to three, trimester-specific time points.

### Estimation of WBC Proportions

Cell type proportions were estimated from the epigenome-wide DNA methylation data using Houseman’s reference-based approach implemented in the ‘estimatecellcounts2’ function from the FlowSorted.Blood.EPIC package in R (Houseman et al., 2012; Salas et al., 2018). An external reference data set of epigenome-wide DNA methylation data generated from pure cell subtypes isolated from blood samples was used as a reference to infer cell type proportions in our sample (Salas et al., 2018). The reference data set was derived from samples obtained from healthy individuals who had no history of significant or chronic health issues and consisted of samples from 31 male and 6 non-pregnant female donors who had a mean age of 32.6 years and average weight of 86 kg (189.2 pounds) (Salas et al., 2018).

To estimate our cell type proportions, we used 450 CpG probes from the EPIC array that were previously identified as being differentially methylated based on cell type by the Identifying Optimal Libraries (IDOL) algorithm (Salas et al., 2018). Of those 450 IDOL probes, DNA methylation data were available for 384 probes that passed our sample QC. Our approach yielded proportion data for six cell types: neutrophils, monocytes, B cells, Natural Killer (NK) cells, CD4+ T cells, and CD8+ T cells. Reference data for basophils and eosinophils were not available and therefore estimation of these cell types was not possible in the current study.

However, proportions and variability of eosinophils and basophils are not typically estimated using this method, presumably as they contribute the least amount of variability in cell type proportions (Adalsteinsson et al., 2012).

### Statistical Analyses

Statistical analyses were conducted using R version 3.6.0 (R Core Team, Vienna, Austria) and SPSS version 23 (IBM, Chicago, IL). Demographic and clinical characteristics were compared between groups using t-tests or Pearson’s chi square test. A p-value of <0.05 was considered statistically significant. To compare cell type proportions between cases and controls, proportion means and standard deviations (SD) were calculated for each cell type within each trimester first for the entire sample, and then stratified by self-reported race. Next, Houseman’s regression calibration approach with a double bootstrap bias estimate was used to formally test differences between cell type proportions in cases versus controls (Houseman et al., 2012). DNA methylation data for 384 IDOL probes were extracted from the external reference data set and our genome-wide DNA methylation data. Using those data, we then used a linear mixed effects model with a random intercept for batch effects to simultaneously estimate all cell type proportion differences by regressing our cell type proportion matrix on case/control status at three cross-sectional, trimester-specific time points. This method employs a double-bootstrap procedure in which both the reference data set and the test data set are sampled with replacement 250 times to compute a standard error estimate used to asses bias due to measurement error, subsequently accounting for variation and noise in cell type proportions estimated from DNA methylation data in small data sets (Houseman et al., 2012). Within each trimester, double bootstrap bias-adjusted regression coefficient estimates, which can be interpreted as differences in cell type proportions (in % points) in cases versus controls, as well as double bootstrap standard errors, 95% confidence intervals (CI), and p-values were computed.

## Results

### Sample Demographics and Clinical Characteristics

Table 1 displays sample demographic and clinical characteristics. Our overall sample consisted of 56 participants (28 cases, 28 controls). After methylation quality control checks, our sample consisted of 24 cases and 26 controls for Trimester 1 and 25 cases and 28 controls for Trimesters 2 and 3. A majority of the participants in the sample self-reported their race as Black (n=42, 75%). By study design, mean (±SD) pre-pregnancy BMI for cases and controls was in the obese range at 33.0 (±7.5) and 33.7 (±7.5), respectively. Between cases and controls, there were no significant differences in maternal age at delivery, pre-pregnancy BMI, gestational age at sample collection, proportion of nulliparity, average systolic blood pressure prior to gestational week 20, and proportion of lifetime smokers. According to the American College of Obstetricians and Gynecologists’ parameters for the trimesters of pregnancy, the time points for mean gestational age place sample collection during the second half of each trimester (American College of Obstetricians and Gynecologists, 2018). Expected significant differences characteristic of the PE phenotype included: gestational age at delivery, average diastolic blood pressure prior to gestation week 20 (although within normal range), average systolic and diastolic blood pressures in labor, and birthweight of infants.

**Table 1.** Demographics and Clinical Characteristics of Sample

| Characteristics | Cases (PE +) | Controls (PE -) | *p* |
| --- | --- | --- | --- |
| *n* | 28 | 28 | ----- |
| *Self-Reported Race: Black Participants* | 21 (75%) | 21 (75%) | ----- |
| *White Participants* | 7 (25%) | 7 (25%) | ----- |
| *Maternal Age at Delivery, years* | 23.94 ± 5.00 | 23.66 ± 4.26 | 0.820<sup>a</sup> |
| *Gestational Age at Delivery, weeks* | 37.38 ± 2.45** | 39.52 ± 1.31 | <0.001<sup>a</sup> |
| *Pre-pregnancy BMI, kg/m<sup>2</sup>* | 32.99 ± 7.49 | 33.68 ± 7.52 | 0.734<sup>a</sup> |
| *Gestational Age at Sample Collection, weeks:* |  |  |  |
| *Trimester 1* | 8.46 ± 1.72 | 8.57 ± 1.75 | 0.817<sup>a</sup> |
| *Trimester 2* | 19.74 ± 1.63 | 19.62 ± 0.79 | 0.752<sup>a</sup> |
| *Trimester 3* | 37.07 ± 2.45 | 37.41 ± 2.48 | 0.612<sup>a</sup> |
| *Nulliparous, n (%): Yes* | 21 (75%) | 24 (85.71%) | 0.313<sup>b</sup> |
| *Average SBP <20 Weeks, mmHg* | 112.79 ± 7.27 | 111.54 ± 8.89 | 0.567<sup>a</sup> |
| *Average DBP <20 Weeks, mmHg* | 70.11 ± 5.60* | 66.82 ± 6.13 | 0.048<sup>a</sup> |
| *Average SBP in Labor, mmHg* | 148.29 ± 6.91** | 124.54 ± 6.06 | <0.001<sup>a</sup> |
| *Average DBP in Labor, mmHg* | 90.54 ± 8.10** | 71.01 ± 6.25 | <0.001<sup>a</sup> |
| *Birthweight (grams)* | 2797 ± 732.79* | 3385.61 ± 540.80 | <0.05<sup>a</sup> |
| *Lifetime Smoking Status, n (%): No* | 18 (62.29%) | 19 (67.86%) | 0.778<sup>b</sup> |

*Note.* Values represent indicated units ± standard deviation. *p*=p-value. *BMI*= body mass index.
*SBP*= systolic blood pressure. *DBP*= diastolic blood pressure. *mmHg*= millimeters of mercury.
*g*= grams. <sup>a</sup>Two sample t-test. <sup>b</sup>Pearson Chi-Square test. \**p* < 0.05. \*\**p* < 0.001.

### Cell Type Proportions Across Pregnancy

Figure 1 displays mean (±SD) proportions estimated for WBCs at Trimesters 1, 2, and 3 for cases and controls. Results are presented for the entire sample (N=56), as well as stratified by self-reported race (Black, n=42; White, n=14). Raw values are provided in Supplemental Table 1. Neutrophils comprised the highest proportion of WBCs at any given time point for both cases and controls, regardless of race (Figure 1A). The remaining five estimated cell types accounted for smaller proportions, with CD4+ T cells representing the second largest proportion, followed by CD8+ T cells, monocytes, B cells, and NK cells. The mean proportions of neutrophils increased significantly across pregnancy from Trimester 1 to Trimester 3 in both cases and controls in the overall sample, as well as in Black and White participant groups separately (Figure 1A). Mean neutrophil proportions increased with each chronological trimester for all groups regardless of case/control status, although not all of these differences were statistically significant. In contrast, monocytes were the most static cell type across pregnancy, with few significant changes by trimester, and no significant differences across pregnancy from Trimester 1 to Trimester 3 (Figure 1B).

**Figure 1.**
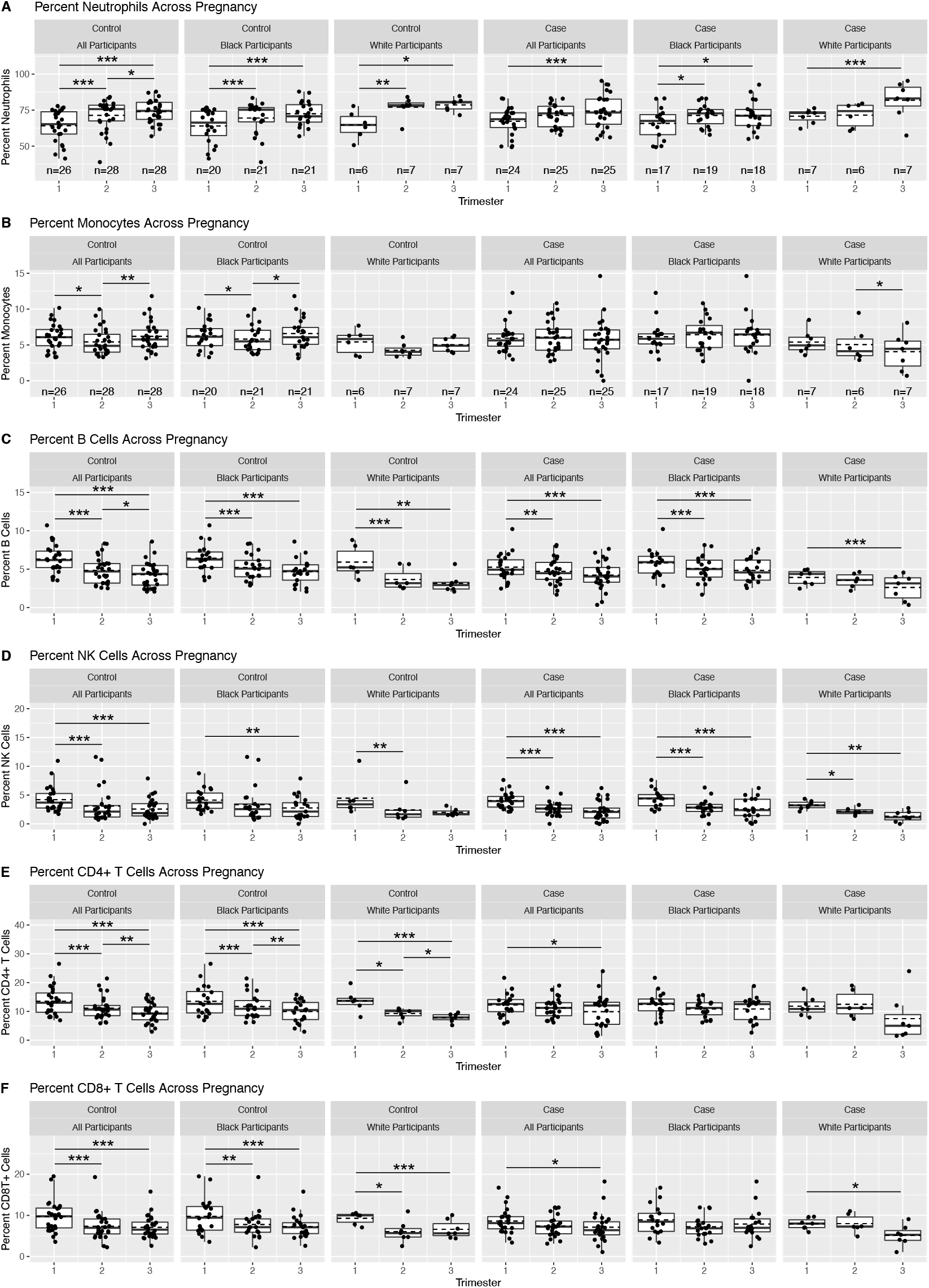
Estimations of WBC Proportions at Trimesters 1, 2, and 3 of Preeclamptic or Normotensive Pregnancy, Overall and Stratified by Self-reported Race. Dots represent proportions for individual participants, represented as percentages; lower and upper hinges correspond to the first and third quartiles (25th and 75th percentiles); dashed, horizontal line indicates the mean; solid, horizontal line indicates the median; whiskers extending from the hinge represent the largest and smallest values no further than 1.5 times the interquartile range; data beyond the whiskers represent outliers; paired t-test: ∗p < 0.05, ∗∗p < 0.005, ∗∗∗p < 0.0005.

Mean proportions of B cells and NK cells declined across pregnancy regardless of case/control status and race, though not every reduction between trimesters achieved statistical significance (Figure 1C-D). A significant reduction in B cell proportions was apparent from Trimester 1 to Trimester 2 in all groups, except within White participant cases (Figure 1C).

Similarly, NK cell proportions decreased significantly across pregnancy (Trimester 1 to Trimester 3) in all groups tested, except within White participant controls, who decreased only from Trimester 1 to Trimester 2 (Figure 1D).

Mean CD4+ T cell proportions decreased significantly across pregnancy in both cases and controls in the overall sample (Figure 1E). Significant reductions were seen in each trimester within both Black and White participant controls, but no meaningful differences were observed within Black or White participants controls. Similarly, mean proportions of CD8+ T cells decreased significantly across pregnancy in the overall sample for both cases and controls (Figure 1F). However, the pattern observed in the overall sample for cases was only evident for White participants in race-stratified results.

### Cell Type Proportions in Cases versus Controls

Figure 2 displays mean (±SD) proportions for estimated WBCs in cases compared to controls at Trimesters 1, 2, and 3. Results are presented for the entire sample (N=56), as well as stratified by self-reported race (Black, n=42; White, n=14). Raw values are provided in Supplemental Table 1. Table 2 presents the double-bootstrap regression coefficient estimates for cases versus controls, double-bootstrap standard error, 95% CI, and p-values for the entire sample, as well as the sample stratified by race. In Trimester 1, within the subset of participants who self-reported their race as White, the mean proportion of neutrophils was significantly higher in cases compared to controls, with an effect size estimate of 5.32% (95% CI = 0.47 to 10.17%, *p* = 0.04) (Figure 2A, Table 2). Interestingly, in Trimester 2, the mean neutrophil proportions in cases compared to controls among White participants were significantly lower, with an estimate of -4.86% (95% CI = -9.14 to −0.58%, *p* = 0.02). There was no significant difference observed in neutrophil proportions for Trimester 3 among White participants. We found no significant differences in the mean proportion of neutrophils between cases and controls in the overall sample or among self-reported Black participants specifically.

**Table 2.** Regression Coefficient Estimates, Representing Differences in Cell Type Proportions, in Percentages, in PE Cases Compared to Controls

| **Trimester 1** |  |  |  |  |  |  |  |  |  |  |  |  |
| --- | --- | --- | --- | --- | --- | --- | --- | --- | --- | --- | --- | --- |
|  | **Overall Sample, n=50** |  |  |  | **Black Participants, n=37** |  |  |  | **White Participants, n=13** |  |  |  |
|  | **Est** | **SE** | **95% CI** | **p** | **Est** | **SE** | **95% CI** | **p** | **Est** | **SE** | **95% CI** | **p** |
| (Intercept, $\gamma_0$) | 0.08 | 0.28 | -0.46 to 0.62 | 0.77 | 0.23 | 0.34 | -0.43 to 0.89 | 0.46 | -0.46 | 0.43 | -1.30 to 0.39 | 0.29 |
| Neutrophils | 2.90 | 2.10 | -1.22 to 7.02 | 0.20 | 1.64 | 2.66 | -3.57 to 6.85 | 0.55 | 5.32 | 2.47 | 0.47 to 10.17 | 0.04* |
| Monocytes | 0.09 | 0.50 | -0.90 to 1.07 | 0.84 | 0.00 | 0.67 | -1.31 to 1.31 | 0.95 | 0.47 | 0.69 | -0.88 to 1.83 | 0.43 |
| B cells | -1.03 | 0.43 | -1.87 to -0.19 | 0.02* | -0.50 | 0.49 | -1.46 to 0.45 | 0.34 | -2.26 | 0.65 | -3.53 to -0.99 | 0.001* |
| NK cells | 0.28 | 0.37 | -0.44 to 0.99 | 0.35 | 0.62 | 0.42 | -0.20 to 1.45 | 0.14 | -0.45 | 0.60 | -1.63 to 0.73 | 0.43 |
| CD4+ T cells | -1.05 | 1.11 | -3.24 to 1.13 | 0.37 | -0.91 | 1.39 | -3.64 to 1.82 | 0.55 | -1.46 | 1.50 | -4.40 to 1.48 | 0.36 |
| CD8+ T cells | -1.16 | 0.79 | -2.70 to 0.38 | 0.16 | -0.96 | 0.98 | -2.87 to 0.96 | 0.31 | -1.02 | 0.53 | -2.06 to 0.01 | 0.05 |
| **Trimester 2** |  |  |  |  |  |  |  |  |  |  |  |  |
|  | **Overall Sample, n=53** |  |  |  | **Black Participants, n=40** |  |  |  | **White Participants, n=13** |  |  |  |
|  | **Est** | **SE** | **95% CI** | **p** | **Est** | **SE** | **95% CI** | **p** | **Est** | **SE** | **95% CI** | **p** |
| (Intercept, $\gamma_0$) | -0.46 | 0.29 | -1.03 to 0.11 | 0.12 | -0.27 | 0.39 | -1.04 to 0.49 | 0.47 | -0.64 | 0.42 | -1.46 to 0.18 | 0.13 |
| Neutrophils | 0.53 | 1.57 | -2.54 to 3.60 | 0.80 | 1.40 | 1.83 | -2.19 to 5.00 | 0.35 | -4.86 | 2.18 | -9.14 to -0.58 | 0.02* |
| Monocytes | 0.60 | 0.45 | -0.28 to 1.49 | 0.19 | 0.52 | 0.52 | -0.51 to 1.55 | 0.31 | 0.91 | 0.68 | -0.42 to 2.23 | 0.17 |
| B cells | -0.07 | 0.31 | -0.69 to 0.54 | 0.85 | -0.07 | 0.34 | -0.74 to 0.59 | 0.65 | -0.21 | 0.33 | -0.86 to 0.43 | 0.51 |
| NK cells | -0.23 | 0.32 | -0.87 to 0.40 | 0.46 | -0.31 | 0.38 | -1.06 to 0.44 | 0.39 | 0.55 | 0.39 | -0.22 to 1.32 | 0.15 |
| CD4+ T cells | -0.22 | 0.76 | -1.71 to 1.27 | 0.89 | -0.89 | 0.94 | -2.74 to 0.96 | 0.27 | 2.53 | 1.32 | -0.06 to 5.12 | 0.06 |
| CD8+ T cells | -0.09 | 0.60 | -1.26 to 1.08 | 0.93 | -0.29 | 0.66 | -1.57 to 0.98 | 0.59 | 1.63 | 0.89 | -0.12 to 3.39 | 0.07 |

| Trimester 3 |  |  |  |  |  |  |  |  |  |  |  |  |
| --- | --- | --- | --- | --- | --- | --- | --- | --- | --- | --- | --- | --- |
|  | Overall Sample, n=53 |  |  |  | Black Participants, n=39 |  |  |  | White Participants, n=14 |  |  |  |
|  | Est | SE | 95% CI | *p* | Est | SE | 95% CI | *p* | Est | SE | 95% CI | *p* |
| (Intercept, $\gamma_0$) | -0.18 | 0.39 | -0.96 to 0.59 | 0.60 | -0.10 | 0.37 | -0.82 to 0.62 | 0.69 | 0.16 | 0.50 | -0.81 to 1.13 | 0.78 |
| Neutrophils | 0.45 | 1.97 | -0.34 to 4.32 | 0.78 | -0.76 | 2.08 | -4.84 to 3.32 | 0.80 | 2.54 | 3.87 | -5.06 to 10.13 | 0.49 |
| Monocytes | -0.73 | 0.56 | -1.82 to 0.36 | 0.17 | -0.88 | 0.71 | -2.26 to 0.51 | 0.21 | -0.27 | 0.73 | -1.71 to 1.17 | 0.68 |
| B cells | 0.14 | 0.36 | -0.57 to 0.85 | 0.72 | 0.42 | 0.41 | -0.38 to 1.23 | 0.36 | -0.63 | 0.34 | -1.29 to 0.03 | 0.06 |
| NK cells | -0.14 | 0.32 | -0.77 to 0.48 | 0.67 | -0.04 | 0.36 | -0.75 to 0.66 | 0.91 | -0.24 | 0.45 | -1.13 to 0.64 | 0.57 |
| CD4+ T cells | 0.64 | 1.02 | -1.36 to 2.63 | 0.51 | 0.95 | 1.00 | -1.02 to 2.92 | 0.40 | -0.12 | 2.89 | -5.79 to 5.55 | 0.95 |
| CD8+ T cells | 0.02 | 0.65 | -1.26 to 1.30 | 0.98 | 0.54 | 0.73 | -0.90 to 1.97 | 0.48 | -1.42 | 0.79 | -2.97 to 0.12 | 0.08 |

*Note.* *Est*= Double-bootstrap regression coefficient estimate. *SE*= Double-bootstrap standard error. *CI*= Confidence Interval. *p*= p-value. \*Significance based on an alpha of 0.05.

**Figure 2.**
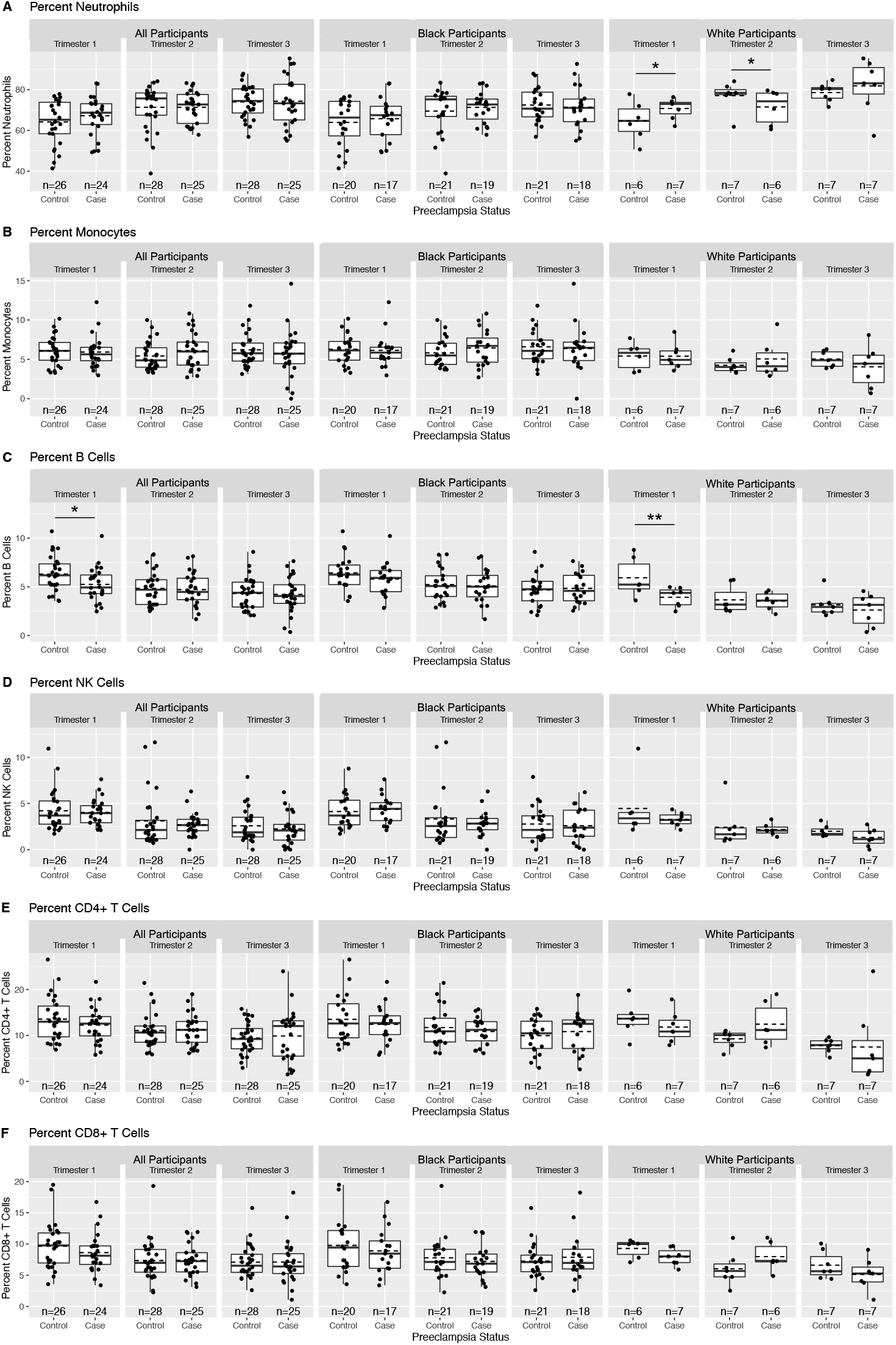
Comparison of WBC Proportion Estimations between Preeclamptic Cases and Normotensive Controls at Trimesters 1, 2, and 3 of Pregnancy, Overall and Stratified by Self-reported Race. Dots represent proportions for individual participants, represented as percentages; lower and upper hinges correspond to the first and third quartiles (25th and 75th percentiles); dashed, horizontal line indicates the mean; solid, horizontal line indicates the median; whiskers extending from the hinge represent the largest and smallest values no further than 1.5 times the interquartile range; data beyond the whiskers represent outliers; double bootstrap: ∗p < 0.05, ∗∗p < 0.005, ∗∗∗p < 0.0005.

In the overall sample in Trimester 1, the mean proportion of B cells was lower in cases compared to controls, with an estimate of -1.03% (95% CI = -1.87 to −0.19, *p* = 0.02) (Figure 2C, Table 2). In the subsample of participants who self-reported their race as White, this difference persisted with an estimate of -2.26 (95% CI = -3.53 to −0.09, *p* = 0.001). In participants who self-reported their race as Black, the proportion of B cells in cases was lower than controls, but not statistically significantly (*p* = 0.34). Group differences in B cell proportions in Trimesters 2 and 3 were not significant in the overall sample, or among results stratified by race. Finally, we found no significant differences in the proportions of monocytes, NK cells, CD4+ T cells, or CD8+ T cells across pregnancy between cases and controls in the overall sample, or in the results stratified by self-reported race across pregnancy (Figure 2B, 2D-F).

## Discussion

We sought to evaluate differences in WBC proportions estimated from epigenome-wide DNA methylation data from individuals who developed PE during pregnancy compared to normotensive controls. We have yet to come across another study evaluating these six WBC types (neutrophils, monocytes, B cells, NK cells, CD4+ T cells, CD8+ T cells) across the three trimesters of preeclamptic and normotensive pregnancy. In our sample, neutrophils represented the largest proportion of the estimated cell types, and our findings demonstrated a steady increase in the mean proportions of neutrophils for both PE cases and controls (Figure 1A, Supplemental Table 1). This finding is consistent with the literature, which suggests that the increase in WBC count observed in pregnancy is due to an increase in the number of neutrophils (Canzoneri, Lewis, Groome, & Wang, 2009; Gabbe, 2017; Gatti et al., 1994). We observed minimal variations in monocytes in both PE cases and normotensive controls (Figure 1B).

Monocytes have previously been shown to remain relatively static across healthy pregnancy (Gabbe, 2017), and no studies were found in which monocytes were measured across preeclamptic pregnancy. We did not come across any comparable studies evaluating longitudinal changes in B cells, NK cells, CD4+ T cells, and CD8+ T cell proportions, or values across preeclamptic or normotensive pregnancy for comparison.

In our analyses comparing PE cases to controls, in the overall sample, we found a lower mean proportion of B cells among cases during Trimester 1 (Figure 2C). Neutrophils, monocytes, NK cells, CD4+ T cells, and CD8+ T cells did not differ between cases and controls at any given time point (Figure 2A-B, 2D-F). In addition to B cells, we estimated proportions for three additional cell types of lymphoid origin: NK cells, CD4+ T cells, and CD8+ T cells.

Previous studies have demonstrated a statistically significant, markedly lower lymphocyte count in PE patients compared to controls in the first trimester and at term (Gabbe, 2017; Kirbas et al., 2015; Lurie et al., 1998). A limitation of these studies is that by measuring all “lymphocytes” they cannot discern which particular cell type(s) are responsible for the reduction. Our data suggest that a lower proportion of B cells may be driving the marked reduction reported in lymphocytes in cases in these studies. In pregnancy, B cells have been shown to produce protective antibodies that are directed toward paternal antigens and associated with positive pregnancy outcomes (Muzzio, Zenclussen, & Jensen, 2013). It is possible that during the first trimester, a reduction in B cells yields insufficient protective antibodies, resulting in a reduction in the immune system’s ‘tolerance’ of the fetus in early pregnancy. These data conflict with the findings reported by Örgül et al. (2019), who found no differences in lymphocytes in the first trimester. Stratifying our results by self-reported race allowed us to further investigate differences in WBC proportions between cases and controls and generated meaningful results. We found B cell proportions to be statistically lower in cases among White participants only, and no statistically significant differences among Black participants. It is possible that our conflicting lymphocyte results are related to differences in the racial demographics of our sample compared to that used by Örgül et al. (2019). Racial breakdown was not provided for comparison.

We found no significant differences in mean neutrophil proportions between cases and controls in our overall sample (Figure 2A; Table 2). Previous investigations exploring differences in neutrophil values between PE cases and normotensive controls have reported conflicting results. Lurie et al. (1998) found neutrophils to be elevated among both mild (n=30) and severe (n=16) PE cases compared to controls (n=46) at the onset of labor. Kirbas et al. (2015) report no differences in neutrophils in neither mild (n=288) nor severe (n=326) PE cases compared to controls (n=320) during the first trimester. In contrast, Örgül et al. (2019) reported elevated neutrophils among both early-onset (n=21) and late-onset (n=42) PE cases compared to controls (n=123), during the first trimester. It should be noted that these studies were performed in samples comprised of individuals with a pre-pregnancy BMI in the normal range, while our sample consisted of mostly obese individuals. In our race-stratified results, we did find a greater proportion of neutrophils in cases in the Trimester 1, as well as a lower proportion of neutrophils in cases in Trimester 2, among White participants only. We did not find statistically significant differences between cases and controls among Black participants. Again, a potential explanation for the discrepancies across the literature and our results may be differences in racial demographics of the samples. Unfortunately, the authors of the three papers discussed above did not provide the racial distribution of their samples for comparison. An additional surprising finding from our study was the significantly lower proportion of neutrophils in cases compared to controls among White participants during Trimester 2 (Figure 1A; Table 2). We have yet to come across another study reporting WBC values or such differences in neutrophils in PE cases during the second trimester. While this difference may reflect the dynamic nature of inflammation and angiogenesis in PE, it is possible that this result is a product of small sample size of White participants (n=14) and this finding must be considered with caution until tested in a larger sample to confirm the result. We found no significant differences in neutrophil proportions between cases and controls among Black participants only. Finally, we found no statistically significant differences in monocyte, NK cell, CD4+ T cell, and CD8+ T cell proportions between cases and controls across pregnancy in our overall results or results stratified by race (Figure 2B, 2D-F; Table 2). Our results for monocytes are consistent with two previous studies characterizing WBC differences in PE relative to normotensive pregnancy by flow cytometry during the third trimester (Melgert et al., 2012; van Nieuwenhoven, Moes, Heineman, Santema, & Faas, 2008). In addition to monocytes, Van Nieuwenhoven et al. (2008) quantified NK cells, CD4+ T cells, and CD8+ T cells and reported no significant differences between PE cases (n=15) compared to controls (n=19) in median total number of cells, which is similar to our results for Trimester 3. They do, however, report a significantly lower median of NK cells as a percentage of lymphocyte population in PE cases, which we did not evaluate in our analyses (van Nieuwenhoven et al., 2008).

### Strengths and Limitations

There are many strengths to this study, including its rigorous design and longitudinal approach. Participants were frequency matched on self-reported race, maternal age at delivery, pre-pregnancy BMI, gestational age at sample collection, nulliparity, average systolic and diastolic blood pressure <20 weeks of gestation, and smoking status, minimizing demographic differences. Stringent clinical criteria were used to precisely define the case and control phenotypes. This is also the first study, to our knowledge, to report WBC proportions estimated from longitudinal DNA methylation data in a study of PE. An additional notable strength of our study was that 75% of our sample self-reported their race as Black. While studies have been conducted to determine differences in complete blood counts in healthy adults by race, we were unable to identify such studies that have been performed in adults during pregnancy (Cheng, Chan, Cembrowski, & van Assendelft, 2004; Lim et al., 2010). Samples used for establishing WBC values in preeclamptic pregnancy have either been collected as part of studies of predominantly White participants, or studies that do not report racial demographics. While establishment of WBC parameters all races is important, it is particularly dire for Black individuals, who have the highest rate of PE, yet continue to be under-represented in studies.

Despite these strengths, there are limitations that should be noted. First, our sample size was small, which may have limited our ability to detect important signals in the data. Our sample of cases also included a mix of late-onset (n=16), early-onset (n=2), and pre-term PE (n=10), which limits the ability to discern any subtype-specific differences that may exist. Finally, our sample exclusively consisted of mainly overweight and obese adults, and it has been established that obesity is associated with chronic inflammation and increased WBC values, which may have reduced the generalizability of this study (Veronelli et al., 2004).

### Conclusion and Future Directions

PE is characterized by chronic inflammation, yet few studies have investigated WBC changes across preeclamptic pregnancy. We capitalized on epigenome-wide DNA methylation data to estimate proportions of neutrophils, monocytes, B cells, NK cells, CD4+ T cells, and CD8+ T cells in PE cases and normotensive controls during the three trimesters. When PE cases were compared to controls, we observed significantly higher neutrophil proportions in Trimester 1 and significantly lower in Trimester 2, as well as significantly lower B cell proportions in Trimester 1 in White participants only (n=14). These findings underscore differences in WBC proportions by race, as no significant findings were observed among Black participants (n=42).

Longitudinal studies with larger sample sizes, representing a population that is racially diverse, includes all BMI categories, and PE subtypes are needed to follow up on our preliminary findings. Furthermore, researchers have begun to further classify PE into molecularly distinct subclasses (Leavey, Bainbridge, & Cox, 2015; Leavey et al., 2016). We speculate that cases of inflammatory PE may be more common among White individuals, as evidenced by their changes in WBC proportions. Similarly, it is possible that different subclasses of PE are more prevalent among Black individuals, as we did not see any significant differences among cases in Black participants in our study. Such trends could be evaluated by performing group trajectory-based analyses in larger samples. Establishment of personalized WBC reference values specific to PE and race could provide healthcare providers with an additional surveillance tool. Such parameters could also provide further insights into PE pathophysiology, including a potential loss of immunomodulation related to B cell insufficiency in early pregnancy.

**Outside of funding supporting this study, the authors declare no conflicts of interest**.

## Data Availability

dbGAP, accession number: phs001937.v1.p1

https://www.ncbi.nlm.nih.gov/gap/advanced_search/?TERM=phs001937.v1.p1

